# Investigating the Impact of Social Determinants of Health on Diagnostic Delays and Access to Antifibrotic Treatment in Idiopathic Pulmonary Fibrosis

**DOI:** 10.1101/2024.11.15.24317403

**Authors:** Rui Li, Qiuhao Lu, Andrew Wen, Jinlian Wang, Sunyang Fu, Xiaoyang Ruan, Liwei Wang, Hongfang Liu

**Affiliations:** McWilliams School of Biomedical Informatics, The University of Texas Health Science Center at Houston, Houston, TX, USA

## Abstract

Idiopathic pulmonary fibrosis (IPF) is a rare disease that is challenging to diagnose. Patients with IPF often spend years awaiting a diagnosis after the onset of initial respiratory symptoms, and only a small percentage receive antifibrotic treatment. In this study, we examine the associations between social determinants of health (SDoH) and two critical factors: time to IPF diagnosis following the onset of initial respiratory symptoms, and whether the patient receives antifibrotic treatment. To approximate individual SDoH characteristics, we extract demographic-specific averages from zip code-level data using the American Community Survey (via the U.S. Census Bureau API). Two classification models are constructed, including logistic regression and XGBoost classification. The results indicate that for time-to-diagnosis, the top three SDoH factors are education, gender, and insurance coverage. Patients with higher education levels and better insurance are more likely to receive a quicker diagnosis, with males having an advantage over females. For antifibrotic treatment, the top three SDoH factors are insurance, gender, and race. Patients with better insurance coverage are more likely to receive antifibrotic treatment, with males and White patients having an advantage over females and patients of other ethnicities. This research may help address disparities in the diagnosis and treatment of IPF related to socioeconomic status.

## Introduction

Idiopathic pulmonary fibrosis (IPF) is a chronic, progressive lung disease characterized by the thickening and scarring (fibrosis) of lung tissue. It is the most common form of pulmonary fibrosis^1^. In the United States, the annual incidence of IPF is estimated to range from 6.8 to 8.8 cases per 100,000 people^2^. Diagnosing IPF is challenging for several reasons: its symptoms are similar to those of other lung diseases, its rarity and gradual progression make early detection difficult, and there are currently no reliable biomarkers. Furthermore, delayed referrals to specialized centers exacerbate the issue, leading to frequent late diagnoses. Before receiving a formal IPF diagnosis, many patients endure years of respiratory symptoms. During this time, they typically undergo numerous examinations and tests to rule out other potential causes of fibrosing interstitial lung disease (ILD)^3^.

Despite the prolonged diagnostic process, another critical concern is treatment. Recent research suggests that antifibrotic therapy may significantly reduce the risks of all-cause mortality, hospitalization, acute exacerbations, and mortality following acute exacerbations in patients with IPF^4,5^. FDA has approved two drugs for the treatment of IPF: nintedanib and pirfenidone^6^. However, researches show that only 10.3% of patients with IPF were treated with an antifibrotic during their disease course^7^.

To improve individual and population health, reduce health disparities, and advance health equity, the healthcare community has increasingly focused on research surrounding social determinants of health (SDoH) in recent years^8^. Studies have demonstrated that SDoH are strongly linked to disease mortality, with unfavorable SDoH profiles independently associated with poorer health outcomes^9,10^. Numerous studies have also explored the relationship between SDoH and idiopathic pulmonary fibrosis (IPF)^11,12^, though they primarily focus on the impact of SDoH on lung transplant eligibility and outcomes. However, given that timely diagnosis and access to antifibrotic treatment are critical for IPF management, an important question arises: how do SDoH influence both the diagnosis and treatment of IPF?

In this paper, we examine the associations between SDoH and two critical concerns, time to IPF diagnosis following initial respiratory symptoms and whether the patient receives antifibrotic treatment, which may further help to mitigate inequalities in the diagnosis and treatment of IPF based on socioeconomic status.

## Methods

### Data Source

We use UTHealth OMOP CDM dataset. It contains EHR data from the outpatient practice of the University of Texas Health Sciences Center at Houston’s McGovern Medical School, and the EHR data is further transformed to the Observational Health Data Sciences and Informatics’ Observational Medical Outcomes Partnership Common Data Model (OMOP CDM).

### Study Population

We included all patients aged 50 years or older at the time of their first clinical diagnosis of idiopathic pulmonary fibrosis (IPF), identified by ICD-9 code 516.31 and ICD-10 code J84.112^7^. The cohort selection process is outlined in Figure 1. A total of 1,221 patients were diagnosed with either ‘J84.112’ or ‘516.31’, of which 1,110 were aged 50 or older at the time of diagnosis. From this group, we excluded patients whose first recorded respiratory symptoms occurred before ‘1900-01-01’, leaving 1,029 patients eligible for analysis of antifibrotic treatment. To study the time from initial recorded respiratory symptoms to IPF diagnosis, we further excluded patients whose respiratory symptoms were recorded after their IPF diagnosis, as the symptoms should precede the diagnosis. Cases where respiratory symptoms were documented after the diagnosis may be due to incomplete electronic health records within the UTHealth OMOP CDM system. Some patients may have experienced symptoms long before visiting UT Physicians, and these earlier records were not captured. Ultimately, 679 patients remained for the analysis of time to IPF diagnosis.

**Figure 1:**
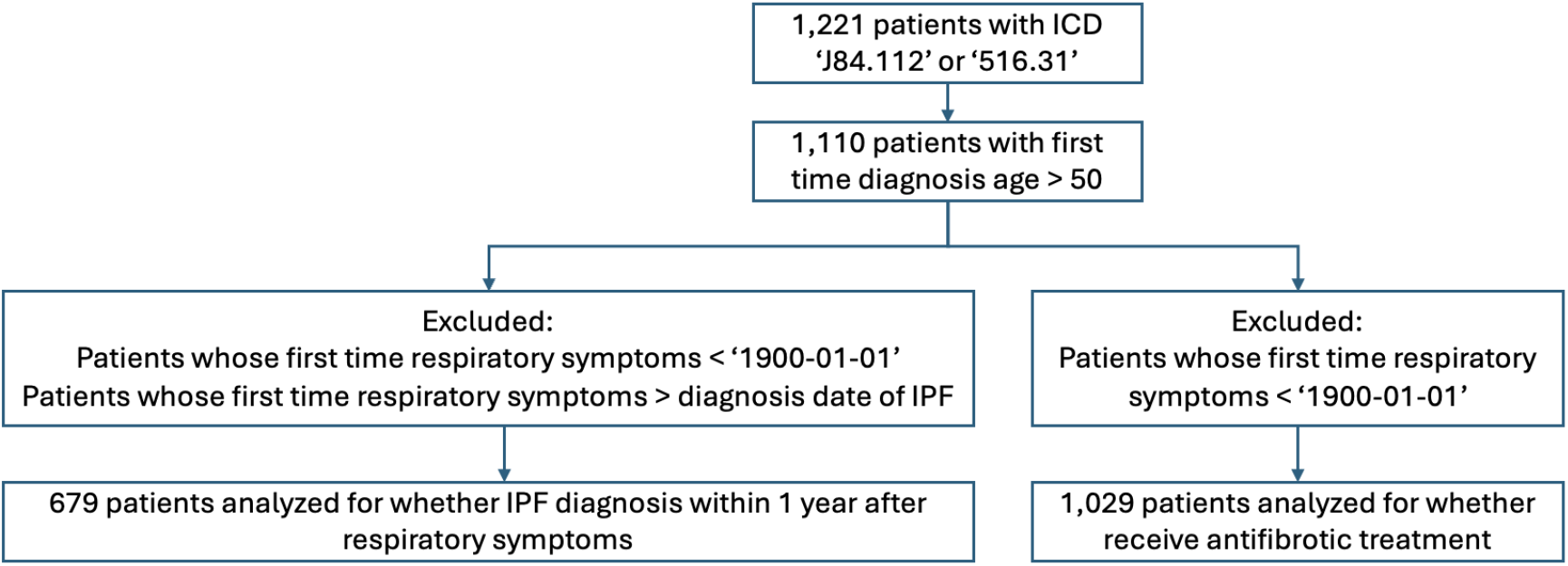
Study Flowchart.

### Ascertainment of Clinical Related Outcomes

We selected two critical clinical outcomes for our analysis: (1) the time to IPF diagnosis after the initial onset of respiratory symptoms, and (2) whether the patient received antifibrotic treatment. Previous research has shown that diagnosing IPF is challenging due to the need to exclude other potential causes of fibrosing interstitial lung disease (ILD)^3,7^. Additionally, studies have revealed that many patients with IPF saw multiple physicians and waited over a year before receiving the correct diagnosis^13^. We framed the time-to-diagnosis problem as a binary classification task: a label of 1 was assigned if the IPF diagnosis occurred within one year of the patient’s first recorded respiratory symptoms, and a label of 0 if the diagnosis took longer than one year. Respiratory symptoms were identified based on the criteria listed in Table 2 of [7]. For the treatment, antifibrotic therapies such as Nintedanib and Pirfenidone have been shown to reduce short-term mortality and hospitalizations in IPF patients^4,14,15^. Therefore, we classified treatment outcomes as a binary classification problem: label 1 indicated that the patient received antifibrotic therapy, while label 0 indicated they did not.

### Ascertainment of SDoHs

We selected the SDoH features based on the five domains defined by the U.S. Department of Health and Human Services (HHS)^16^. These domains include Economic Stability, Education Access and Quality, Health Care Access and Quality, Neighborhood and Built Environment, and Social and Community Context. For this analysis, four key SDoH features were chosen: household income (reflecting economic stability), the percentage of individuals with a bachelor’s degree or higher (representing education access and quality), health insurance status (indicating health care access), and PM2.5 levels (serving as a proxy for neighborhood and built environment). Since the dataset in UTHealth OMOP CDM lacks direct information on household income, education, and insurance status, we utilized the patient’s zip code, age, gender and race to extract area-specific averages from the American Community Survey, accessed via the U.S. Census Bureau’s API^17^. These demographic-specific averages were then assigned to each patient as representative SDoH data. For PM2.5 data, we converted zip codes to Federal Information Processing Standards (FIPS) codes and retrieved air quality information using the Air Quality System (AQS) API from the U.S. Environmental Protection Agency^18^. Among the available PM2.5 metrics, we selected the quarterly nighttime mean PM2.5 levels for our analysis. Additionally, following the World Health Organization’s SDoH framework, which includes Socioeconomic and Political Contexts as upstream factors, and Ethnicity (Racism) as a Socioeconomic Position factor^19,20^, we incorporated demographic variables such as age at first diagnosis of IPF, gender, and race to further account for these critical dimensions.

### Statistical Analysis

As previously mentioned, both clinical related outcomes are formulated as binary classification tasks. To investigate the associations between SDoH and clinical outcomes, we begin by building machine learning models that take SDoH features as input and output a binary label corresponding to clinical outcomes. Using these models, we then compute and interpret the associations between the input features and the outcomes. In the following section, we first introduce the selected classification models, followed by a detailed explanation of how we analyze the association between the input features and the outcome labels.

The model inputs consist of both numerical features, such as diagnosis age, household income, education level, insurance status, and PM2.5 levels, as well as categorical features including gender and race. Numerical features are standardized, while categorical features are transformed using one-hot encoding, which creates new binary columns for each category. To analyze the associations from different perspectives, we selected two commonly used classification models: logistic regression and XGBoost. To validate the effectiveness of two classification models, we evaluate the classification performance using the following three metrics:

- **F1 score**: *F* 1 = 2(*Pre · Rec*)*/*(*Pre* + *Rec*), where *Pre* is precision and *Rec* is recall.
- **Average Precision**: *AP* = ∑_*n*_(*Rec*_*n*_ − *Rec*_*n*−1_) *Pre*_*n*_, it computes the weighted sum of precision, with the increase in recall from the previous threshold used as the weight, where *Pre*_*n*_ and *Rec*_*n*_ are the precision and recall at the *n*-th threshold. It reflects the trade-off between precision and recall across thresholds.
- **Cohen’s Kappa**: *κ* = (*p*_*o*_ − *p*_*e*_)*/*(1 − *p*_*e*_), where *p*_*o*_ is the observed aggrement, which is identical to accuracy, and *p*_*e*_ is the expected aggrement, which is probabilities of randomly seeing each category.

Logistic regression is a fundamental classification algorithm defined as 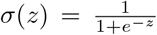, where *z* = *β*_0_ + *β*_1_*X*_1_ + *β*_2_*X*2 + … + *β*_*n*_*X*_*n*_. *β*_0_, *β*_1_, …, *β*_*n*_ are the model coefficients, and *X*_1_, *X*_2_,, *X*_*n*_ are the input features. The coefficients *β*_1_, …, *β*_*n*_ quantify the impact of each feature on the log-odds of the outcome: a positive coefficient increases the log-odds, while a negative coefficient decreases it. The magnitude of the coefficient reflects the strength of the feature’s influence on the outcome, with larger absolute values indicating stronger influence. We also compute the odds ratio, defined as *odds ratio* = *e*^*β*^, which quantifies how the odds of the outcome change with a one-unit increase in the corresponding feature. By comparing the coefficients and odds ratios across different SDoH features, we can assess the relative importance of each feature in predicting the outcome.

XGBoost classification is an efficient model based on the gradient boosting mechanism. It constructs an ensemble of decision trees in a sequential manner, where each subsequent tree focuses on correcting the errors made by the previous ones. Compared to logistic regression, XGBoost typically achieves better performance with early stopping and built-in regularization. But it lacks direct interpretability in terms of coefficients. To address this, we compute the relative risk (RR) to compare the probability of predicting the positive class (label = 1) across different cohort groups. Relative risk is defined as 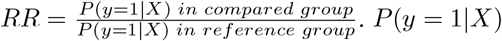 represents the probability that the XGBoost model predicts the label as 1, given the input SDoH features X. If *RR >* 1, the probability of predicting label 1 is higher in the comparison group than in the reference group. Conversely, if *RR <* 1, the probability is lower. For numerical features, we discretize them into ordinal categories by grouping continuous values into ranges and assigning a category to each range. We then select the smallest range as the reference group and compute the relative risk for other ranges. This approach enables us to examine the relationships between different ranges of SDoH features and the outcome, offering valuable insights into how variations within a single SDoH feature impact the predicted probabilities.

## Experiments

### Dataset Statistic

Out of the 1,110 patients diagnosed with IPF at age 50 or older, 1,045 reside in Texas. Figure 2 shows the county-level distribution of these patients within Texas. The color intensity represents the number of patients in each county; darker shades indicate a higher number of patients. Notably, the majority of patients are concentrated in the Houston area, with Harris County having the highest number at 669.

**Figure 2:**
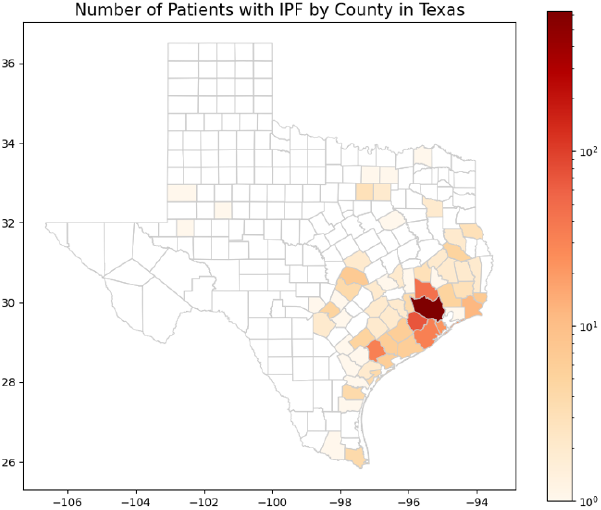
The County-level Distribution of Patients with IPF in Texas.

Figure 3 further illustrates trends in time-to-diagnosis and the percentage of patients receiving antifibrotic treatment over time. The x-axis represents the year, with the left y-axis showing the percentage, and the right y-axis indicating the total number of patients. Figure 3 (a) shows the distribution of time from the first recorded respiratory symptoms to IPF diagnosis across different years, based on 679 patients analyzed for the time-to-diagnosis study. Darker blue bars represent patients diagnosed within one year of initial respiratory symptoms, while lighter blue bars represent those diagnosed after more than one year. We can observe that the percentage of patients diagnosed within one year decreases from 2015 to 2019 and remains relatively stable between 2020 to 2024. In 2015, all patients were diagnosed within one year, possibly due to incomplete EHR data from UT Physicians, as some patients may have experienced symptoms before their first visit. The total number of patients increased steadily from 2015 to 2022 but declined between 2022 and 2024. Figure 3 (b) depicts the trends in whether patients received antifibrotic treatment over the years, based on 1,029 patients included in the antifibrotic treatment analysis. Darker blue bars represent patients who received antifibrotic treatment. The data indicates that patients began receiving antifibrotic treatment after 2015, with the percentage increasing from 2015 to 2022, followed by a decline after 2020. The total number of patients grew from 2011 to 2022, and start to decrease between 2022 and 2024.

**Figure 3:**
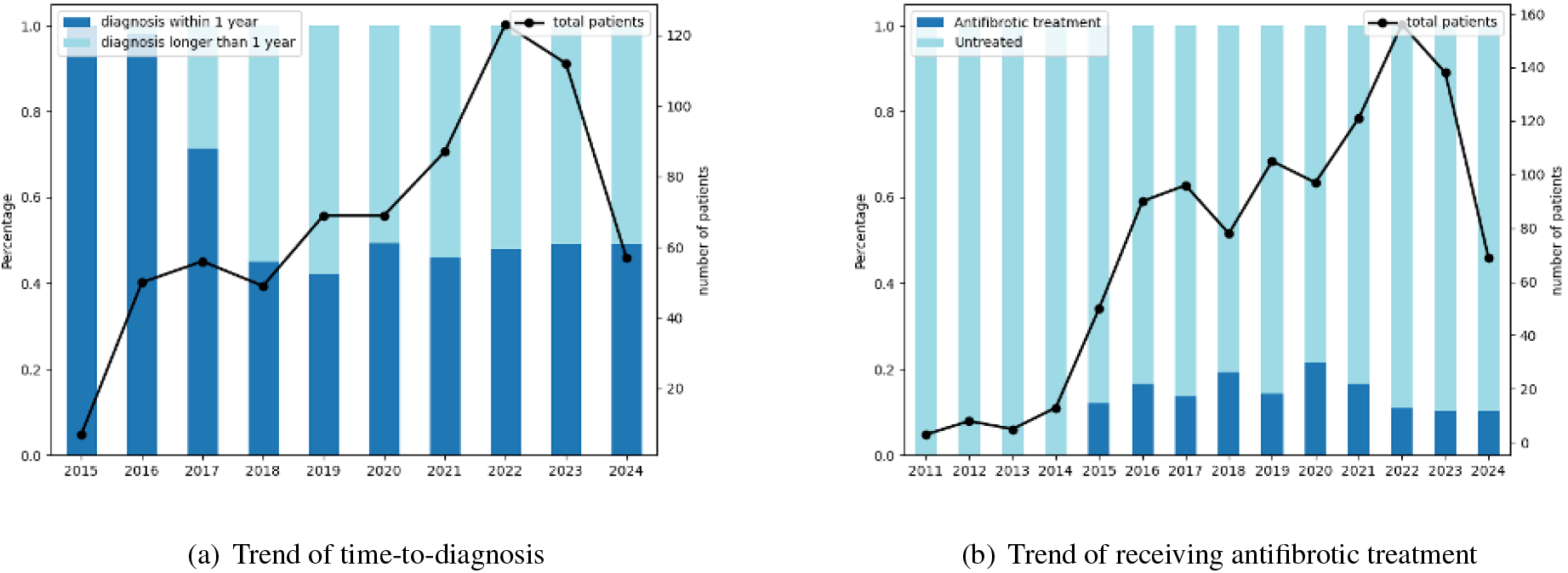
Trends of Treatment/Diagnosis Time

Table 1 shows the demographics and SDoH for studied patients. Because the dataset from UTHealth OMOP CDM does not include income, education, or insurance information, we extracted demographic-specific averages from zip code-level data provided by the American Community Survey through the U.S. Census Bureau’s API^17^. The average SDoH features for patients in the same area, race, gender, and age group were used as proxies for individual patient data. The dataset covers 315 distinct zip codes, though 10 zip codes were missing from the census API. Additionally, 26 patients had missing income and insurance information, and 29 had missing education data. For these missing values, we used the average value from the available data for imputation.

**Table 1:**
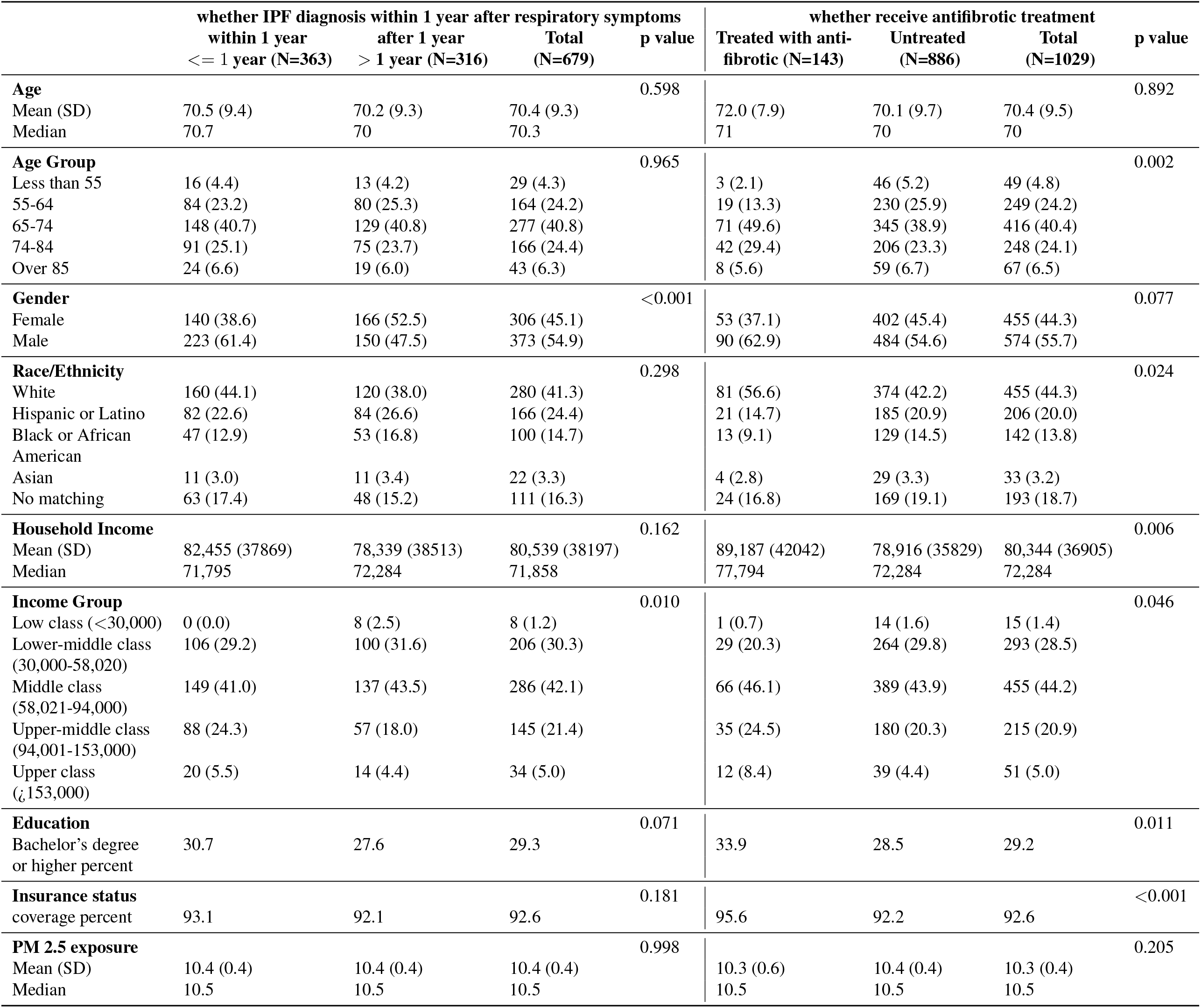
Demographics and SDoH of Studied Patients.

**Table 2:** Classification Performance.

|  | whether IPF diagnosis within 1 year<br>after respiratory symptoms |  | whether receive anti-<br>fibrotic treatment |  |
| --- | --- | --- | --- | --- |
|  | logistic regression | XGBoost classification | logistic regression | XGBoost classification |
| F1 | 0.605 | 0.600 | 0.268 | 0.250 |
| AP | 0.510 | 0.619 | 0.177 | 0.262 |
| Cohen's Kappa | 0.107 | 0.109 | 0.064 | 0.187 |

For the analysis of whether IPF diagnosis occurred within one year of initial respiratory symptoms, 363 patients were diagnosed within one year, while 316 were diagnosed after one year. We calculated the statistics for both groups, with p-values used to compare their distributions. A p-value less than 0.05 was considered statistically significant. There was no significant difference in the age of first diagnosis between the two groups. However, in terms of gender, males were more likely to receive a faster diagnosis than females. No statistically significant differences were found for race or income between the two groups. However, when household income was categorized into ordinal classes, a higher proportion of patients in the upper-middle to upper-class income brackets were diagnosed within one year. Additionally, there were no significant differences between the two groups in terms of education, insurance status, or PM2.5 exposure. For the analysis of antifibrotic treatment, the distribution was more imbalanced, with 143 patients receiving antifibrotic treatment and 886 patients remaining untreated. This finding is consistent with previous research^7^, which reported that only 10.4% of patients received antifibrotic treatment. Although there was no significant difference in the age of first diagnosis, when age was categorized into ordinal groups, a higher proportion of patients aged between 65–84 were more likely to receive antifibrotic treatment. There was no significant difference in gender. However, white patients were more likely to receive treatment compared to other ethnicities. Patients with higher income and education levels were also more likely to receive antifibrotic treatment. No statistically significant difference was found for PM2.5 exposure.

### Classification Performance

For both studies, we split the dataset into training and test sets using an 85:15 ratio. To address class imbalance, we adjusted the sample weights to assign more importance to the minority class. Table 2 presents the classification performance on the test set. Both tasks show that the models achieve fair performance, with particularly better results in the time-to-diagnosis task, demonstrating the effectiveness of our classification approach. Better performance could be achieved by incorporating patient-level SDoH features rather than relying on demographic averages from zip code-level data, which only serve as proxies for individual patient characteristics. Comparing the two models, the XGBoost classifier outperforms logistic regression in terms of average precision (AP) and Cohen’s Kappa, although it yields a slightly lower F1 score. The higher AP indicates that XGBoost predicts fewer false positives, maintaining precision even as recall increases, and excels at identifying positive cases. Meanwhile, a higher Cohen’s Kappa shows that XGBoost handles class imbalance more effectively and achieves stronger agreement with the true labels. However, the slightly lower F1 score suggests that while XGBoost reduces false positives, it may miss some true positives.

### Association between SDoHs and Clinical Outcomes

We first compute and analyze the associations between SDoH and clinical outcomes for logistic regression model, which naturally provides interpretability through its coefficients. The magnitude of each coefficient reflects the strength of a feature’s influence on the outcome, with larger absolute values indicating stronger effects. A positive coefficient increases the log-odds of the outcome, while a negative coefficient decreases it. Table 3 presents the coefficients alongside their corresponding odds ratios. Categorical features were encoded using one-hot encoding, converting each category into a separate binary column. Our analysis shows that the top three SDoH factors most strongly associated with the time-to-diagnosis task are education, gender, and insurance. Specifically, patients with higher educational attainment and better insurance coverage are more likely to receive a diagnosis within one year of their initial symptoms. Males tend to receive quicker diagnoses compared to females. For antifibrotic treatment, the most influential SDoH features are insurance status, race, and gender. Patients with insurance are significantly more likely to receive antifibrotic treatment. Males are more likely to receive the treatment compared to females, and White patients are more likely to be treated than other racial groups.

**Table 3:** Associations between SDoH and Clinical Outcomes in Logistic Regression.

|  | whether IPF diagnosis within 1 year<br>after respiratory symptoms |  | whether receive anti-<br>fibrotic treatment |  |
| --- | --- | --- | --- | --- |
|  | Coefficient | Odds Ratio | Coefficient | Odds Ratio |
| age | -0.089 | 0.915 | 0.021 | 1.021 |
| income | -0.199 | 0.819 | 0.062 | 1.064 |
| education | 0.310 | 1.363 | 0.007 | 1.007 |
| insurance | 0.174 | 1.190 | 0.331 | 1.392 |
| PM2.5.mean | -0.182 | 0.833 | -0.1 | 0.905 |
| **Gender** |  |  |  |  |
| Female | -0.309 | 0.734 | -0.169 | 0.844 |
| Male | 0.309 | 1.363 | 0.170 | 1.185 |
| **Race** |  |  |  |  |
| White | -0.075 | 0.927 | 0.285 | 1.330 |
| Hispanic or Latino | -0.003 | 0.997 | -0.240 | 0.787 |
| Black or African American | -0.046 | 0.955 | -0.049 | 0.952 |
| Asian | -0.222 | 0.801 | 0.057 | 1.059 |
| No matching concept | 0.347 | 1.415 | 0.053 | 0.948 |

After comparing the feature importance among various SDoH features, we further examine how variations within each individual SDoH feature impact the predicted probabilities. Table 4 presents the relative risk ratios for XGBoost classification models given 95% CI. Numerical features are discretized into ordinal categories, and we select a range as the reference group. Relative risks for other ranges are then calculated accordingly. Education, insurance, and PM2.5 levels are grouped into two categories: low and high. For age at first diagnosis, we use the 65-74 age group as the reference. As age increases, patients are generally more likely to receive a quicker diagnosis; however, this trend reverses for patients aged 85 and older, where the probability decreases. For gender, with females as the reference group, males are more likely to receive a quicker diagnosis. In terms of race, Black or African American patients serve as the reference group, and the model shows that White patients tend to receive quicker diagnoses. When considering household income, middle class is chosen as the reference group, and patients with higher incomes show a greater likelihood of receiving quicker diagnoses. Additionally, individuals with higher levels of education and with insurance are more likely to experience quicker diagnoses. Finally, for PM2.5 exposure, patients living in areas with lower PM2.5 levels are more likely to receive a quicker diagnosis.

**Table 4:** Associations between SDoHs and Clinical Outcomes in XGBoost Classification.

|  | whether IPF diagnosis within 1 year<br>after respiratory symptoms | whether receive anti-<br>fibrotic treatment |
| --- | --- | --- |
|  | risk ratio (95% CI) | risk ratio (95% CI) |
| **Age Group** |  |  |
| Less than 55 | 0.628 (0.169-1.349) | 0.136 (0.028-0.326) |
| 55-64 | 0.709 (0.529-0.934) | 0.553 (0.111-1.365) |
| 65-74 | 1.000 (reference) | 1.000 (reference) |
| 74-84 | 1.091 (0.887-1.331) | 1.314 (0.465-2.846) |
| Over 85 | 0.706 (0.372-1.096) | 0.762 (0.048-2.451) |
| **Gender** |  |  |
| Female | 1.000 (reference) | 1.000 (reference) |
| Male | 1.477 (1.205-1.832) | 1.766 (0.701-4.576) |
| **Race/Ethnicity** |  |  |
| White | 1.054 (0.818-1.399) | 4.849 (1.028-22.948) |
| Hispanic or Latino | 0.795 (0.545-1.093) | 1.281 (0.187-5.644) |
| Black or African<br>American | 1.000 (reference) | 1.000 (reference) |
| Asian | 0.648 (0.515-0.851) | 2.102 (0.0167-10.626) |
| no matching | 0.942 (0.651-1.289) | 1.749 (0.089-10.039) |
| **Income Group** |  |  |
| Low class (<30,000) | 0.121 (0.091-0.156) | 0.078 (0.022-0.182) |
| Lower-middle class<br>(30,000-58,020) | 0.858 (0.681-1.065) | 0.809 (0.198-1.961) |
| Middle class<br>(58,021-94,000) | 1.000 (reference) | 1.000 (reference) |
| Upper-middle class<br>(94,001-153,000) | 1.343 (1.094-1.627) | 2.362 (0.764-5.787) |
| Upper class<br>(>153,000) | 1.121 (0.769-1.542) | 4.553 (0.166-11.686) |
| **Education** |  |  |
| low education level | 1.000 (reference) | 1.000 (reference) |
| high education level | 1.233 (1.024-1.471) | 1.908 (0.751-4.315) |
| **Insurance status** |  |  |
| low insurance level | 1.000 (reference) | 1.000 (reference) |
| high insurance level | 1.291 (1.056-1.575) | 1.696 (0.724-3.508) |
| **PM 2.5 status** |  |  |
| low PM2.5 level | 1.000 (reference) | 1.000 (reference) |
| high PM2.5 level | 0.818 (0.684-0.981) | 0.743 (0.333-1.541) |

## Discussion

In this study, we examine the associations between social determinants of health (SDoH) and clinical outcomes for patients with idiopathic pulmonary fibrosis (IPF) at UT Physicians. We focus on four SDoH variables: household income, education level, insurance status, and PM2.5 exposure. Additionally, demographic factors such as age at first diagnosis, gender, and race are included in the analysis. Since the UTHealth OMOP CDM dataset lacks direct records for income, education, insurance, and PM2.5 exposure, we use zip code-level data from the American Community Survey (U.S. Census Bureau API^17^) as proxies for individual SDoH characteristics. Two key clinical outcomes are investigated: time to IPF diagnosis following initial respiratory symptoms and whether the patient receives antifibrotic treatment. The average time to a diagnosis of IPF was 642 days (1.75 years), which was shorter than 2.7 years in the previous research^7^. This may be due to the incomplete EHR in UTHealth OMOP CDM dataset. And about 13.9% patients were treated with antifibrotics. This finding is consistent with previous research^7^, which reported that only 10.4% of patients received antifibrotic treatment. Both clinical outcomes are modeled as binary classification tasks, employing logistic regression and XGBoost classifiers. For logistic regression, we calculate coefficients and odds ratios to assess the impact of SDoH variables on the clinical outcomes. For XGBoost, relative risk ratios are computed to evaluate how variations within each SDoH feature affect predicted probabilities. Our findings indicate that gender, education, insurance status, and race are the most influential SDoH variables associated with the two outcomes. Specifically, males tend to receive quicker diagnoses and are more likely to receive antifibrotic treatment compared to females. Patients with higher educational attainment and insurance coverage also experience faster diagnoses and greater likelihood of receiving antifibrotic treatment. Additionally, higher household income correlates with shorter time to diagnosis and higher probability of receiving treatment.

Our contribution can be summarized in the following two aspects: (1) We extract demographic-specific averages from zip code-level data using the American Community Survey (via the U.S. Census Bureau API) as proxies for individual SDoH characteristics. Unlike composite measures such as the Area Deprivation Index (ADI), our approach allows for a more fine-grained investigation of the associations between individual SDoH features and clinical outcomes. (2) We build two machine learning models—logistic regression and XGBoost classification—to examine the impact of SDoH variables on clinical outcomes. In the logistic regression model, we assess and compare the feature importance of SDoH variables using coefficients, while in the XGBoost classification model, we compute relative risk ratios to evaluate how variations in each SDoH feature influence predicted probabilities.

Our study has the following two limitations. (1) Limited Cohort Size: The cohort analyzed in this study is relatively small, with only 1,029 patients in the analysis of antifibrotic treatment and 679 patients in the analysis of whether IPF diagnosis occurred within one year of initial respiratory symptoms. The small sample size may reduce the generalizability of our findings to larger populations. (2) Single-Registry Data: Our analysis is based solely on data from UTHealth OMOP CDM, a single registry. This could introduce bias, and we cannot confirm whether our conclusions would hold consistently across other registries or broader patient populations. Besides, some patients may have experienced symptoms long before visiting UT Physicians, and these earlier records were not captured. In the future, we plan to extend our research by analyzing and comparing the associations between SDoH features and clinical outcomes using datasets from multiple registries to enhance the robustness and generalizability of our findings.

## Data Availability

All data produced in the present study are available upon reasonable request to the authors

## Acknowledgements

This project is supported by the Cancer Prevention Research Institute of Texas (CPRIT)RR230020, and National Human Genome Research Institute R01HG12748.

## References

1. American Lung Association. Idiopathic Pulmonary Fibrosis (IPF); 2024. Available from: https://www.lung.org/lung-health-diseases/lung-disease-lookup/idiopathic-pulmonary-fibrosis#:~:text=Idiopathic%20pulmonary%20fibrosis%20(IPF)%20is,makes%20it%20difficult%20to%20breathe.

2. Nalysnyk L, Cid-Ruzafa J, Rotella P, Esser D. Incidence and prevalence of idiopathic pulmonary fibrosis: review of the literature. European Respiratory Review. 2012;21(126):355–61.

3. Ryu JH, Moua T, Daniels CE, Hartman TE, Eunhee SY, Utz JP, et al. Idiopathic pulmonary fibrosis: evolving concepts. In: Mayo Clinic Proceedings. vol. 89. Elsevier; 2014. p. 1130–42.

4. Dempsey TM, Sangaralingham LR, Yao X, Sanghavi D, Shah ND, Limper AH. Clinical effectiveness of antifibrotic medications for idiopathic pulmonary fibrosis. American journal of respiratory and critical care medicine. 2019;200(2):168–74.

5. Kang J, Han M, Song JW. Antifibrotic treatment improves clinical outcomes in patients with idiopathic pulmonary fibrosis: a propensity score matching analysis. Scientific reports. 2020;10(1):15620.

6. American Lung Association. Pulmonary Fibrosis Medications; 2024. Available from: https://www.lung.org/lung-health-diseases/lung-disease-lookup/pulmonary-fibrosis/patients/how-is-pulmonary-fibrosis-treated/medications.

7. Herberts MB, Teague TT, Thao V, Sangaralingham LR, Henk HJ, Hovde KT, et al. Idiopathic pulmonary fibrosis in the United States: time to diagnosis and treatment. BMC Pulmonary Medicine. 2023;23(1):281.

8. National Institute of Nursing Research. NIH-wide Social Determinants of Health Research Coordinating Committee; 2024. Available from: https://www.ninr.nih.gov/research/nih-sdohrcc.

9. Satti DI, Chan JSK, Dee EC, Lee YHA, Wai AKC, Dani SS, et al. Associations between social determinants of health and cardiovascular health of US adult cancer survivors. Cardio Oncology. 2024;6(3):439–50.

10. Zhao Y, Dimou A, Fogarty ZC, Jiang J, Liu H, Wong WB, et al. Real-world Trends, Rural-urban Differences, and Socioeconomic Disparities in Utilization of Narrow versus Broad Next-generation Sequencing Panels. Cancer Research Communications. 2024;4(2):303–11.

11. Swaminathan AC, Hellkamp AS, Neely ML, Bender S, Paoletti L, White ES, et al. Disparities in lung transplant among patients with idiopathic pulmonary fibrosis: an analysis of the IPF-PRO Registry. Annals of the American Thoracic Society. 2022;19(6):981–90.

12. DeDent AM, Collard HR, Thakur N. Neighborhood health and outcomes in idiopathic pulmonary fibrosis. Annals of the American Thoracic Society. 2024;21(3):402–10.

13. Collard HR, Tino G, Noble PW, Shreve MA, Michaels M, Carlson B, et al. Patient experiences with pulmonary fibrosis. Respiratory medicine. 2007;101(6):1350–4.

14. Kelly BT, Thao V, Dempsey TM, Sangaralingham LR, Payne SR, Teague TT, et al. Outcomes for hospitalized patients with idiopathic pulmonary fibrosis treated with antifibrotic medications. BMC Pulmonary Medicine. 2021;21:1–14.

15. Behr J, Prasse A, Wirtz H, Koschel D, Pittrow D, Held M, et al. Survival and course of lung function in the presence or absence of antifibrotic treatment in patients with idiopathic pulmonary fibrosis: long-term results of the INSIGHTS-IPF registry. European Respiratory Journal. 2020;56(2).

16. Department of Health and Human Services. Social Determinants of Health; 2023. Available from: https://health.gov/healthypeople/priority-areas/social-determinants-health.

17. Census Bureau’s API. American Community Survey 1-Year Supplemental Data; 2022. Available from: https://www.census.gov/data/developers/data-sets/ACS-supplemental-data.html.

18. Environmental Protection Agency. Air Quality System (AQS) API; 2023. Available from: https://aqs.epa.gov/aqsweb/documents/data_api.html#lists.

19. Organization WH, et al. Health 2020: A European policy framework and strategy for the 21st century. World Health Organization. Regional Office for Europe; 2013.

20. Hill-Briggs F, Fitzpatrick SL. Overview of social determinants of health in the development of diabetes. Diabetes Care. 2023;46(9):1590–8.

